# Twitter Discourse on Nicotine as Potential Prophylactic or Therapeutic for COVID-19

**DOI:** 10.1101/2021.01.05.21249284

**Authors:** Ramakanth Kavuluru, Jiho Noh, Shyanika W. Rose

**Affiliations:** Division of Biomedical Informatics, Internal Medicine, 230E MDS Bldg, 725 Rose St, Lexington KY 40506; Computer Science Department, Lexington, KY; Center for Health Equity Transformation and Department of Behavioral Science, College of Medicine, Lexington, KY

## Abstract

**Background:** An unproven “nicotine hypothesis” that indicates nicotine’s therapeutic potential for COVID-19 has been proposed in recent literature. This study is about Twitter posts that misinterpret this hypothesis to make baseless claims about benefits of smoking and vaping in the context of COVID-19. We quantify the presence of such misinformation and characterize the tweeters who post such messages.

**Methods:** Twitter premium API was used to download tweets (n = 17,533) that match terms indicating (a) nicotine or vaping themes, (b) a prophylactic or therapeutic effect, **and** (c) COVID-19 (January-July 2020) as a conjunctive query. A constraint on the length of the span of text containing the terms in the tweets allowed us to focus on those that convey the therapeutic intent. We hand-annotated these filtered tweets and built a classifier that identifies tweets that extrapolate the nicotine hypothesis to smoking/vaping with a positive predictive value of 85%. We analyzed the frequently used terms in author bios, top Web links, and hashtags of such tweets.

**Results:** 21% of our filtered COVID-19 tweets indicate a vaping or smoking-based prevention/treatment narrative. Qualitative analyses show a variety of ways therapeutic claims are being made and tweeter bios reveal pre-existing notions of positive stances toward vaping.

**Conclusion:** The social media landscape is a double-edged sword in tobacco communication. Although it increases information reach, consumers can also be subject to confirmation bias when exposed to inadvertent or deliberate framing of scientific discourse that may border on misinformation. This calls for circumspection and additional planning in countering such narratives as the COVID-19 pandemic continues to ravage our world. Our results also serve as a cautionary tale in how social media can be leveraged to spread misleading information about tobacco products in the wake of pandemics.

## INTRODUCTION

The link between nicotine and COVID-19 has garnered significant attention in both scientific and news media communities. The low prevalence of current smokers among COVID-19 patients and immunomodulatory effects of nicotine are cited as evidence for the claimed therapeutic potential of nicotine for affected patients (Farsalinos, Barbouni, & Niaura, 2020). Although current smokers were found to be at a reduced risk of SARS-CoV-2 infection, meta-analyses reveal that former smokers were at an increased risk of hospitalization, severity, and mortality due to COVID-19 (Simons, Shahab, Brown, & Perski, 2020) while no such associations were found with current smoking status. However, there are several significant limitations of using electronic health record derived determination of smoking status (including missing data on smoking status and lack of adjustment for the demographic profile of those tested/admitted to the hospital), and no studies included in the meta-analysis verified smoking status with biochemical measures. There is growing evidence, on the other hand, that smoking is associated with worse outcomes (hospitalization, death, progression) for COVID-19 patients including two recent thorough meta-analyses (Lowe, Zein, Hatipoğlu, & Attaway, 2021; Patanavanich & Glantz, 2020; Reddy et al., 2021; Zhang et al., 2021).

There is at least one clinical trial planned by French researchers to assess nicotine’s therapeutic and prophylactic potential (Changeux, Amoura, Rey, & Miyara, 2020). However, recently, it has been reported that the first author of the paper discussing the trial has had tobacco industry funding in the past and there are methodological issues in papers indicating the prophylactic role of nicotine (van Westen-Lagerweij et al., 2021). At least one other 2020 paper that has hypothesized the beneficial role of nicotine for COVID-19 has been retracted due to prior links of authors to the tobacco industry (Horel & Keyzer, 2021). Given the rapidly evolving landscape of multiple waves of the epidemic and associated analyses with methodological variations, some researchers conclude that it is too early to conclusively determine nicotine’s role (Edwards & Munafò, 2020).

Overall, in current literature on these themes, authors clearly indicate that smoking’s severe addiction and proven harmful health effects dictate that it shouldn’t be used as a means to prevent/treat COVID-19 Scientific explorations are ongoing about nicotine and its role as a potential COVID-19 therapeutic in controlled settings (Tizabi, Getachew, Copeland, & Aschner, 2020). In the meantime, it is important to understand how this so called “nicotine hypothesis” plays out in consumer discourse on social media regarding beliefs and attitudes towards tobacco products (in the context of the ongoing pandemic). This is especially important in the general context of mistrust of experts and scientific discourse in social media platforms (Baron & Berinsky, 2019). Though Americans in general trust scientists as a career group (National Academies of Sciences & Medicine, 2017), in biomedicine, it appears that the trust is more aligned with practicing physicians as opposed to biomedical researchers (Funk, Hefferon, Kennedy, & Johnson, 2019). However, given COVID-19 is the first “post-truth” epidemic (Parmet & Paul, 2020), the evolving nature of evidence surrounding tobacco products leaves more space for speculation on social media that often goes unchecked. It is well-known that pro-vaping content dominates social media and it has been observed that e-cigarette information seekers on these websites are subsequently more likely to use them (Yang, Liu, Lochbuehler, & Hornik, 2019). Our prior work in this field estimates that nearly 50% of e-cigarette chatter involves marketing themes (Han & Kavuluru, 2016) and identifies tweeters who tweet exclusively pro-vaping content (Kavuluru & Sabbir, 2016). These pro-vaping or smoking posts in the context of COVID-19 may drive more people toward the corresponding products, which is a clear detriment to public health if they are never-users.

In general, health misinformation on social media has been a major issue in the recent years. Two systematic reviews feature tobacco products in the top five categories for health misinformation on social media (Suarez-Lledo & Alvarez-Galvez, 2021; Wang, McKee, Torbica, & Stuckler, 2019). Popular types of misinformation on social platforms surrounding tobacco products include unverified consequences of product use and cessation methods lacking evidence (Tan & Bigman, 2020). Experiments indicate that exposure to videos with misleading information on e-cigarettes and hookah products resulted in viewers having more positive attitudes toward them compared with watching control videos (Albarracin, Romer, Jones, Jamieson, & Jamieson, 2018). Finally, network analyses show that pro-smoking Twitter users form more cohesive networks (via follower-friend links) but also appear to follow anti-smoking tweeters to keep up with regulation and research updates (Park, 2020). Pro-smoking accounts tend to be of individuals while anti-smoking accounts are often associated with organizations (Park, 2020). The increased cohesion creates an opportune environment for pro-smoking tweeters to collectively respond (via replies/comments) to regular tweeters interacting with them to seek information on tobacco products.

With this context in mind, in this paper, we examine publicly available tweets that discuss nicotine or vaping as preventative or treatment options for COVID-19 with associated extrapolations to smoking/vaping’s therapeutic potential in retrieved messages. To our knowledge, there are only two recent efforts that seemed to have touched upon the themes of our study. The first offers an anecdotal indication of unsubstantiated health claims (Majmundar, Allem, Cruz, & Unger, 2020) and does not focus on the nicotine hypothesis. Specifically, three **unsubstantiated** claims were stated: (1) vaping devices can prevent COVID-19 given their ability to increase humidity in lungs, (2) vaping devices can help deliver organic oregano oil to lungs to kill coronavirus, and (3) propylene glycol (often found in e-liquids) can kill airborne COVID-19 particles. The second effort examines sentiment surrounding vaping and smoking in the context of the pandemic (Kamiński, Muth, & Bogdański, 2020); this latter effort’s focus on sentiment has a different purpose of general perceptions of people and not about quantifying misinformation involving vaping/smoking. During the four-month period (January-April 2020), the authors downloaded public timeline tweets mentioning smoking and COVID-19 related terms from a set of tweeters who tweeted on the theme toward the end of April 2020. Even though sentiment was negative in general, it turned relatively less negative during April 2020 when preprints suggesting the preventative role of nicotine surfaced. Our effort has a deliberate focus on misinterpretations of the nicotine hypothesis for COVID-19 spanning tweets from a seven-month duration (January-July 2020). More specifically,

1. We estimate the prevalence of extrapolations of the nicotine hypothesis for COVID-19 to smoking/vaping among the tweets that discuss a beneficial signal of nicotine products. We qualitatively examine the nature of various themes in such tweets with examples of full tweets and top Web links and hash tags.
2. By mining user-specific Twitter bios, we study the thematic predispositions of tweeters who generate extrapolating tweets juxtaposed with those of all tweeters discussing nicotine products in the context of COVID-19.

In a sense, our effort could be construed as a retrospective analysis of what transpired during the first seven months of the pandemic when observational studies were popping up as preprints indicating nicotine’s prophylactic role in covid-19; also, as a cautionary tale on how such preprints were picked up by pro-vaping/smoking tweeters to fit their narrative by extrapolating the already dubious nicotine hypothesis to smoking/vaping.

## METHODS

Using the Twitter Premium API (v1.1) subscription, we downloaded ALL publicly available tweets authored between Jan 1, 2020 and July 31, 2020 containing **at least one term** (or associated hashtag) from each of the three following groups:

1. nicotine, e-cigarette(s), ecig(s), vaping;^*^
2. prevent(s), prevented, preventing, prevention, preventative, preventive, prophylactic, prophylaxis, precaution(ary), treat(s), treated, treating, treatment, cure(s), cured, curing;
3. covid, coronavirus, covid19, corona, covid_19, covid-19, SARS-Cov-2.

We focused on having at least one term from each group because we wanted to explore discussions that are portraying nicotine and associated products as helpful in dealing with COVID-19. We had to use the paid Premium subscription service to launch this query because it is over 300 characters, while the free service only allows a maximum query length of 128 characters. Furthermore, the paid service allowed us to obtain expanded full URLs when tweeters use link shortening services such as *bit.ly*. This was important to rank popular Web pages used in the tweets.

Before we proceed, we note that the IRB at the University of Kentucky deemed that this type of research does not meet the definition of human subjects and thus does not require additional IRB review based on these two criteria: 1. The data is publicly available; and 2. There is no interaction or intervention with subjects. Nevertheless, we only report aggregate metrics (e.g., counts, proportions) and show very few example tweets without author attributions that convey some viewpoints.

Our conjunctive Twitter query outlined earlier in this section led to a total of 17,533 tweets of which 16,064 were in English. We observed several of these tweets were discussing vaping bleach (misinformation propagated by a radio talk show host) for treating COVID-19 and hence we removed all tweets containing “bleach”. This further pruned the dataset to 13,718 tweets. Among these, we imposed a strict constraint that all three terms (one each from three groups above) must occur within a window of six consecutive words (three additional words on top of the three required terms) after commonly used stops words^†^ are removed from the tweet. This was needed to weed out tweets that contain the three terms but are separated by many other terms and whose overall meaning is something quite different from our search intent. For example, consider the tweet by a public health organization in Oregon (USA): “Smoking and **vaping** involve hand-to-mouth contact that may make it easier to spread **COVID-19** to the user and other surfaces. **Prevent** the spread of **COVID-19**, quit today.” Although a term from each group in our query appears here, they are farther apart and do not convey the intent of our search. The six consecutive word window constraint removes this tweet from our data as it does not satisfy it. This window constraint resulted in a smaller subset of 5192 tweets, which is henceforth called the filtered dataset.

From this filtered subset, we (first and second author together) annotated a set of 400 tweets randomly selected to identify if the tweets are actually discussing potential prevention or treatment aspects of nicotine for COVID-19. Among such tweets, we additionally identified those tweets that were extrapolating the nicotine hypothesis to vaping or smoking with direct or implicit claims about vaping or smoking being beneficial in the context of COVID-19. That is, we identified tweets that interpret the nicotine hypothesis to mean beneficiary effects of smoking or vaping. Out of these 400 annotated tweets, 337 turned out to be conveying a treatment/preventative aspect leading to a positive predictive value (PPV) of 84.25% with 95% confidence interval [80.22%, 87.60%].

A supervised machine learned model (Liu, Hsu, & Ma, 1998) was built and evaluated using cross-validation (using the same set of 400 annotated tweets) to identify these tweets that resort to this type of extrapolation based on tweet text and associated user name. The specific model (Liu et al., 1998) is based on association rules that link presence of certain key words to classes (extrapolating tweet or the negative class). The classifier had a positive predictive value (PPV) of 85% with 65% sensitivity. When it was applied to the filtered dataset, 576 were identified as such, translating to over 11% (576/5192) of the dataset. This proportion is smaller than the proportion observed in the human annotated sample because the classifier’s sensitivity is only 65%. We preferred the classifier to be more focused on high precision (PPV) because we wanted to look at the bio text of the Twitter profiles that tweet favorably, involving smoking or vaping. Outside that purpose of separating bios of extrapolating tweeters, the classifier was not thus applied for deriving general proportions.

We ranked top 20 frequent words in the personal bios of Twitter users who tweeted the 576 extrapolating messages (identified by the classifier) as opposed to tweeters in the rest of the tweets. This is to see if tweeters who post such messages had pre-existing viewpoints that may have led them to extrapolate nicotine research to benefits due to smoking or vaping. We also looked at top (most frequent) Web links and hashtags used in extrapolating tweets and the remaining tweets. This was done by just counting the tweets containing each of the unique URLs and hashtags in the dataset.

## RESULTS

Of the 337 tweets (out of 400 hand-annotated messages) identified to be conveying a treatment/preventative aspect, we found 84 to be linking nicotine with smoking/vaping as an extrapolation of research surrounding nicotine or (more rarely) discussing smoking/vaping directly as being therapeutic. This translates to 84/400 = 21% (95% confidence interval: [17.18%, 24.39%]) tweets making such claims. Nearly 80% of these claims involve smoking. Some pertinent examples from our dataset are shown in Table 1 demonstrating the variety of takes on the controversial topic. We only displayed examples here where the tweet contents cannot be linked back to a real person by searching on Twitter. In one instance, a tweeter with over 10,000 followers wondered if vaping was the only thing that can cure COVID-19. In another case, a physician tweeter (with over 2000 followers) from a prestigious U.S. Carnegie R1 university states the facts appearing in a linked news article about the French nicotine clinical trial but also includes a “smoking protects” phrasing in the message that is unwarranted and prone to misinterpretation. Another doctoral level researcher (with over 5000 followers) holding a scientific role in a non-profit organization for smoking cessation takes a nuanced approach that does not outright claim vaping-driven prevention conclusively, but still indicates that there exists evidence supporting it.

**Table 1:**
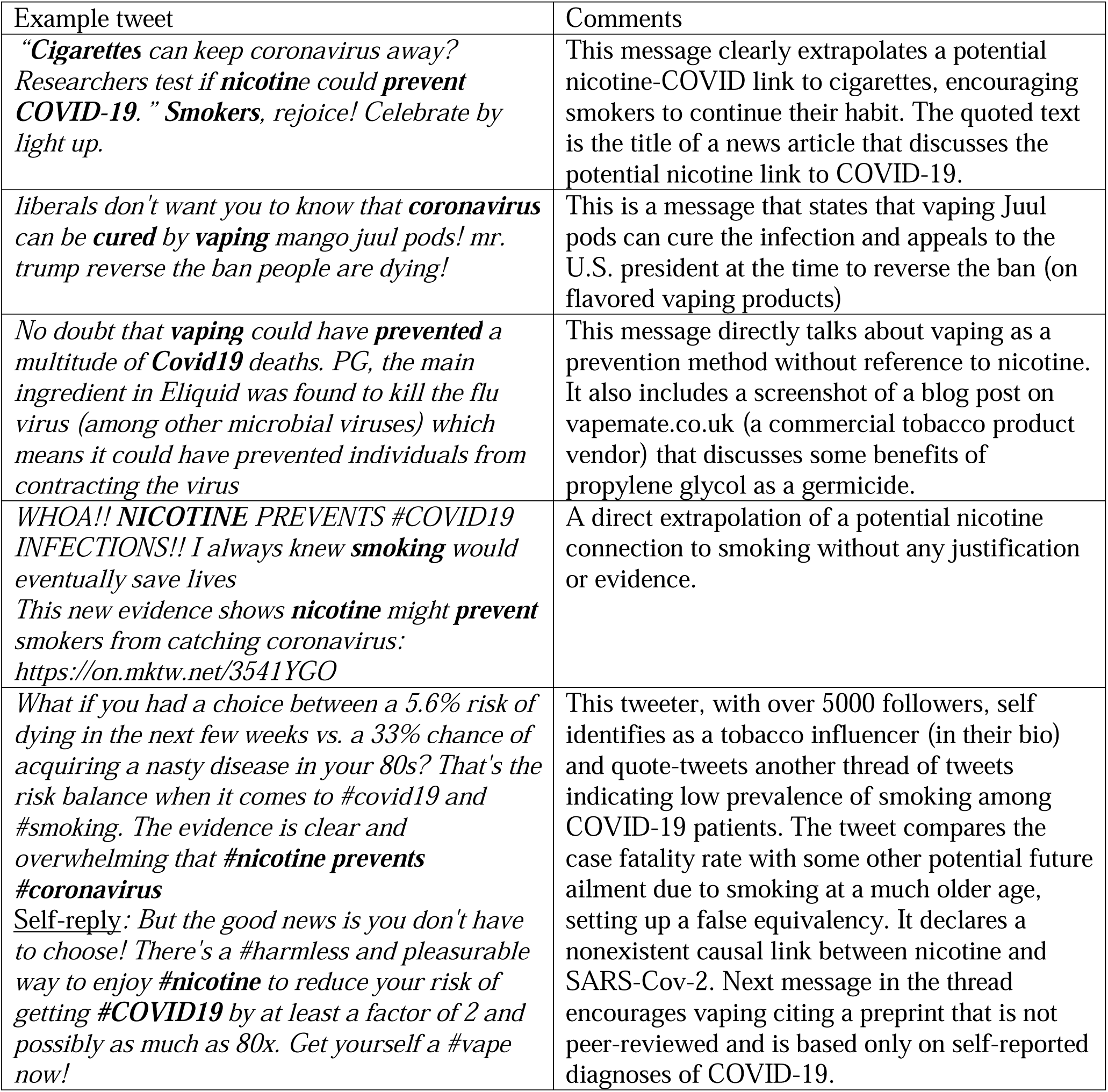
Tweets that appear to indicate smoking or vaping as being protective for COVID-19. Note: Usernames (and display names) corresponding to messages in the table (and the rest of the paper) can only be traced back to personalities not using their real name (e.g., @JohnnyVapes).

| Example tweet | Comments |
| --- | --- |
| *“Cigarettes can keep coronavirus away? Researchers test if **nicotine** could **prevent COVID-19**.” Smokers, rejoice! Celebrate by light up.* | This message clearly extrapolates a potential nicotine-COVID link to cigarettes, encouraging smokers to continue their habit. The quoted text is the title of a news article that discusses the potential nicotine link to COVID-19. |
| *liberals don't want you to know that **coronavirus** can be **cured** by **vaping** mango juul pods! mr. trump reverse the ban people are dying!* | This is a message that states that vaping Juul pods can cure the infection and appeals to the U.S. president at the time to reverse the ban (on flavored vaping products) |
| *No doubt that **vaping** could have **prevented** a multitude of **Covid19** deaths. PG, the main ingredient in Eliquid was found to kill the flu virus (among other microbial viruses) which means it could have prevented individuals from contracting the virus* | This message directly talks about vaping as a prevention method without reference to nicotine. It also includes a screenshot of a blog post on vapemate.co.uk (a commercial tobacco product vendor) that discusses some benefits of propylene glycol as a germicide. |
| *WHOA!! **NICOTINE PREVENTS #COVID19 INFECTIONS!!** I always knew **smoking** would eventually save lives*<br>*This new evidence shows **nicotine** might **prevent** smokers from catching coronavirus:*<br>*https://on.mktw.net/3541YGO* | A direct extrapolation of a potential nicotine connection to smoking without any justification or evidence. |
| *What if you had a choice between a 5.6% risk of dying in the next few weeks vs. a 33% chance of acquiring a nasty disease in your 80s? That's the risk balance when it comes to #covid19 and #smoking. The evidence is clear and overwhelming that **#nicotine prevents #coronavirus***<br>*<u>Self-reply:</u> But the good news is you don't have to choose! There's a #harmless and pleasurable way to enjoy **#nicotine** to reduce your risk of getting **#COVID19** by at least a factor of 2 and possibly as much as 80x. Get yourself a #vape now!* | This tweeter, with over 5000 followers, self identifies as a tobacco influencer (in their bio) and quote-tweets another thread of tweets indicating low prevalence of smoking among COVID-19 patients. The tweet compares the case fatality rate with some other potential future ailment due to smoking at a much older age, setting up a false equivalency. It declares a nonexistent causal link between nicotine and SARS-Cov-2. Next message in the thread encourages vaping citing a preprint that is not peer-reviewed and is based only on self-reported diagnoses of COVID-19. |

Bios are brief descriptions that Twitter users post on their profiles typically talking about themselves. Table 2 shows a distribution of top bio words that occur in profiles of tweeters of the 576 messages that were classified as expressing a favorable view of vaping or smoking for COVID-19 (column 1) in juxtaposition with those from authors of remaining tweets (column 2). The words in the table are sorted in descending order in terms of numbers of bios in that group containing them. The percentages in the parentheses in the table correspond to the proportion of unique tweeters whose bio contains the term. A sample bio of one of the authors reads: “#Vaping ends #smoking harms. Advocate #HarmReduction. Stop the #VapeBan. Deflate nannycrat pseudoscience masquerading as ‘‘#publichealth’’ #AbolishFDA”. It is quite evident from Table 2 that “advocate” and “vaping” feature at the very top in the bios of such authors. Words “advocate” and “vaping” show up in 1.7% and 1.6% of bios of authors in the *other* group, while featuring in 6.3% and 5.7% of author bios in favorable view group, respectively, clearly indicating over 3*X* increase in the latter group. Relatively speaking, the table shows that harm reduction terms are more prominent in the extrapolation tweeter group.

**Table 2:** Top 20 bio words sorted by proportion of unique Twitter users authoring the tweets belonging to each group (extrapolating vs rest); **bold** words are tobacco product related.

| Top bio words for authors of tweets that appear to promote a favorable view of smoking or vaping for COVID-19 | Top bio words for authors of remaining tweets satisfying the window constraint |
| --- | --- |
| Unique tweeters: 508 | Unique tweeters: 4045 |
| **Advocate** (6.3%)<br>**Vaping** (5.7%)<br>**Harm** (3.9%)<br>Love (3.7%)<br>News (3.5%)<br>**Reduction** (3.3%)<br>Trump (3.1%)<br>**Tobacco** (3.1%)<br>**Free** (3.1%)<br>People (2.8%)<br>MAGA (2.6%)<br>Lover (2.6%)<br>Writer (2.6%)<br>Fan (2.4%)<br>Since (2.4%)<br>Media (2.4%)<br>**Smoking** (2.2%)<br>**Vaper** (2.2%)<br>**Smoke** (2.2%)<br>Truth (2.2%) | Love (4.4%)<br>News (3.3%)<br>Life (3.3%)<br>Health (2.5%)<br>World (2.2%)<br>Like (2.2%)<br>Views (2.2%)<br>Fan (2.1%)<br>Tweets (2.1%)<br>Music (2.0%)<br>**Free** (1.9%)<br>Politics (1.8%)<br>Follow (1.8%)<br>People (1.8%)<br>**Advocate** (1.7%)<br>Lover (1.7%)<br>Father (1.6%)<br>Writer (1.6%)<br>**Tobacco** (1.6%)<br>**Vaping** (1.6%) |

We also considered a different way of analyzing tweeters generating the vaping/smoking messages. We begin with a simple definition that any tweeter bio or username containing the following terms as substrings is “pro-vaping”: *smokefree, “smoke free”, “harm reduction”, harmreduction, e-cigarette, ecigarette, vape, vapor, vapour, vaping, ecig, eliquid, ejuice, e-liquid, tobaccofree, tobacco-free, “tobacco free”, thr* (acronym for tobacco harm reduction). Note that because we use substrings, bios containing terms such as *vape*r, #We*Vape*WeVote, #*Vaping*SavesLives, #*Vaping*Advocate will all match our condition. With this definition, 6.8% of tweeters in the filtered dataset are pro-vaping but they generated a total of 12.6% of its tweets. They also author 21.2% of the 576 smoking/vaping tweets.

## DISCUSSION

Our analysis showed that over one in five tweets that talks about the nicotine hypothesis also extrapolates it to a potential benefit of smoking or vaping for COVID-19. Examples from Table 1 and other paraphrased instances show the variety of such tweets ranging from indirect/subtle hints to more explicit declarations. When our classifier was run on unlabeled filtered dataset to identify all such tweets (along with those in the annotated dataset), the corresponding bios reveal vaping related terms featuring at the top (from Table 2). Additionally, results based on the pro-vaping tweeter definition showed that despite constituting less than 7% of tweeters, they authored over 20% of smoking/vaping tweets. This leads us to believe that the tweets implying the potential therapeutic benefits of nicotine (smoked as tobacco or vaped) on COVID-19 are authored by those having pre-existing positive views on vaping or smoking. But, there is potential for casual Twitter users to be exposed to these types of messages providing potentially biased interpretations of scientific findings. This might encourage them to continue their tobacco product consumption habits or initiate new habits, both possibilities that may lead to negative consequences for public health overall. The first column of Table 2 also shows an identification with the current president of the U.S. (Trump) and his campaign slogan (MAGA) potentially indicating associations with political affiliations.

The most frequently used Web link in the favorable group is a news media piece titled: “This new evidence shows nicotine might prevent smokers from contracting the coronavirus” making an indirect link to smoking. The top link in the other group is a news piece titled: “France testing whether nicotine could prevent coronavirus”. It is straightforward to see this link is positing more of a hypothesis while the former one claims evidence why smokers may be protected from the virus. Among the top ten links in both groups (extrapolating vs rest), there are only two links that directly refer to published peer-reviewed research articles (Farsalinos, Barbouni, & Niaura, 2020; Tindle, Newhouse, & Freiberg, 2020) both in the favorable view group.

Upon examining the top ten frequently used hashtags in both groups, we did not find much difference except for one interesting tag #QuitLying that only shows up in the favorable view tweets. This hashtag is often used in replying to tweets by other tweeters who may be tweeting about the potential risks of COVID-19 for smokers and vapers. For instance, consider this tweet: “There’s compelling evidence that #nicotine inhalation prevents #COVID19 infection and reduces the severity of symptoms. Whatever you’re #smoking, it’s clearly addled your brain. #QuitLying”. This was used as a reply to another tweet by a verified medical doctor who tweeted: “Thanks to everyone who joined the @voxmedia, @TobaccoFreeKids + @Bloombergdotorg panel on tobacco use and #COVID19. With over 5M high school students using e-cigarettes, there’s never been a more urgent moment to talk about the connection between tobacco use and the virus”. This hashtag appears to be a mechanism to engage with others who might be having a non-favorable perspective of tobacco products or nicotine for COVID-19.

Before we conclude, we outline some limitations: only public English language tweets were analyzed; so the findings cannot be generalized to the entire conversation about COVID-19 and tobacco use on all social networks especially on platforms like Facebook and WhatsApp where most of the messaging is not public. The discussion is likely evolving as the pandemic changes, and this only captures discussion during the specific duration of January to July in 2020. As the pandemic wanes, the chatter regarding these themes naturally decreases; however, it may still prevail in the highly cohesive networks of pro-smoking/vaping tweeters even if the reach has been curtailed to those outside such networks^‡^. Even if the rate of tweeting on this topic has decreased, we believe it is critical to record the types of misinformation that spread during the peaks of the pandemic and how they can be tracked (using automated classifiers like the one we built in our current effort.)

Overall, our results identify over 5000 tweets that promote the not-yet-validated nicotine hypothesis for COVID-19, which at times is also used to attribute benefits to vaping and smoking, spreading false hope to tobacco product consumers without conclusive evidence. Also given evidence of increased risk of adverse outcomes from COVID-19 among smokers compared with non-smokers, social media messages conflating the ‘nicotine hypothesis’ with potential benefits of smoking for COVID-19 may be detrimental to smokers (Farsalinos, Barbouni, Poulas, et al., 2020). On Twitter, studies have shown that false news stories reach significantly faster and farther than true ones (Vosoughi, Roy, & Aral, 2018). Recent findings also show that exposure to misinformation that smoking helps with COVID-19 actually leads to an increase in tobacco consumption (Luk et al., 2020). Hence, federal health agencies, healthcare organizations, physicians should all be extremely vigilant in communicating verified information to the general public and patients in the wake of nicotine related misinformation about COVID-19. Additionally, engaging credible voices and approaches to provide balanced information and combat misinformation about vaping and smoking and COVID-19 to the public are needed as trust of typical sources of information (e.g., FDA) may also be weakened through this discourse (Freckelton, 2020). On a positive note, studies in health communication have shown that it is possible to correct online misinformation both algorithmically and manually by providing accurate sources of information (Bode & Vraga, 2018; Bode, Vraga, & Tully, 2020; Vraga & Bode, 2018). A future direction for us is to build machine learned models that can spot misinformation and formulate appropriate responses (e.g., as “reply” tweets) with links to scientific evidence.

## Data Availability

The raw tweet content data cannot be shared publicly as per Twitter's policies. In the interest of privacy, we only report aggregate metrics in the manuscript.

## Highlights

- Social media plays an increasingly critical role in tobacco product related communication of both objectively assessed information and biased disinformation.
- Low prevalence of smokers among hospitalized COVID-19 patients is leading to a rapidly evolving scientific discourse on the role of nicotine for COVID-19. It is unclear how this scientific landscape is being interpreted and communicated on social media.
- This paper demonstrates that a nontrivial portion of chatter surrounding nicotine’s role in COVID-19 extrapolates an unproven nicotine hypothesis to vaping and smoking.
- This favorable view of vaping/smoking appears to be mostly carried out by players with pre-existing positive stances regarding the role of vaping in tobacco prevention.
- Our findings warrant vigilance on part of public health agencies and healthcare professionals as they communicate accurate and objective findings at the intersection of nicotine and COVID-19 to counter disinformation.

## Acknowledgments

This work is primarily supported by the U.S. National Cancer Institute (NCI) through NIH grant R21CA218231. Partial support is also provided by the U.S. NCI through grant P30CA177558. The content is solely the responsibility of the authors and does not necessarily represent the official views of the NIH.

## Footnotes

* Most of the sample tweets we downloaded that exclusively mentioned smoking or cigarettes without references to nicotine are genuinely highlighting the dangers of smoking during COVID-19. In light of this and cost effectiveness of our Twitter API subscription to obtain an exhaustive dataset of tweets, we left out “smoking”, “cigarettes”, and “tobacco”. Our goal was to study the chatter surrounding the nicotine mentions.

† http://ir.dcs.gla.ac.uk/resources/linguistic_utils/stop_words

‡ https://help.twitter.com/en/rules-and-policies/twitter-reach-limited

